# Optimization of nutritional strategies using a mechanistic computational model in prediabetes: Application to the J-DOIT1 study data

**DOI:** 10.1101/2023.05.30.23290761

**Authors:** Julia H. Chen, Momoko Fukasawa, Naoki Sakane, Akiko Suganuma, Hideshi Kuzuya, Shikhar Pandey, Paul D’Alessandro, Sai Phanindra Venkatapurapu, Gaurav Dwivedi

## Abstract

Lifestyle interventions have been shown to prevent or delay the onset of diabetes; however, inter-individual variability in responses to such interventions makes lifestyle recommendations challenging. We analyzed the Japan Diabetes Outcome Intervention Trial-1 (J-DOIT1) study data using a previously published mechanistic simulation model of type 2 diabetes onset and progression to understand the causes of inter-individual variability and to optimize dietary intervention strategies at an individual level. J-DOIT1, a large-scale lifestyle randomized intervention study, involved 2607 subjects with a 4.2-year median follow-up period. We selected 112 individuals from the J-DOIT1 study and calibrated the mechanistic model to each participant’s body weight and HbA1c time courses. We evaluated the relationship of physiological (e.g., insulin sensitivity) and lifestyle (e.g., dietary intake) parameters with variability in outcome. Finally, we used simulation analyses to predict individually optimized diets for weight reduction. The model predicted individual body weight and HbA1c time courses with a mean (±SD) prediction error of 1.0 kg (±1.2) and 0.14% (±0.18), respectively. Individuals with the most and least improved biomarkers showed no significant differences in model-estimated energy balance. A wide range of weight changes was observed for similar model-estimated caloric changes, indicating that caloric balance alone may not be a good predictor of body weight. The model suggests that a set of optimal diets exists to achieve a defined weight reduction, and this set of diets is unique to each individual. Our diabetes model can simulate changes in body weight and glycemic control as a result of lifestyle interventions. Moreover, this model could help dieticians and physicians to optimize personalized nutritional strategies according to their patients’ goals.

## Introduction

In the National Diabetes Statistics Report 2020 [1] from the Centers for Disease Control and Prevention (CDC), it was estimated that about 34.2 million people (∼10.5% of the US population) are diabetic, accounting for $237 billion in direct medical expenses and $90 billion in indirect medical costs. Globally, diabetes is now considered an epidemic, affecting more than 420 million individuals (∼6% of the world’s population) [2] and can lead to various complications [3]. Although lifestyle factors, such as diet composition, exercise, and sleep, play an important role in type 2 diabetes (T2D) development [4–6], the response to similar lifestyle changes varies dramatically among individuals [7]. This inter-individual variability could be due to pathophysiological differences among individuals [8], differences in the physiological response to dietary or exercise intervention [9], and other factors [7]. Therefore, it is desirable to develop a framework for designing individualized strategies to achieve defined health goals targeted toward preventing or delaying the onset of diabetes. However, a limited understanding of the causes of inter-individual variability makes it challenging to design individualized interventions, e.g., diet plans, for diabetes prevention.

Precision nutrition aims to prevent and manage chronic diseases by tailoring dietary interventions or recommendations considering the individual’s genetic background, metabolic profile, gut microbiome, and environmental exposure. Currently, the field of precision nutrition is faced with challenges such as the high cost of genomics and metabolomics technologies, and lack robust and reproducible results in studies on precision nutrition [10,11]. In contrast to precision nutrition, there are general strategies that do no attempt to individualize dietary recommendations, such as low-carbohydrate or low-fat diets. Several studies have shown the effectiveness of both low-fat and low-carbohydrate diets for weight control and reduction of cardiovascular risk [12–16]. The US Diabetes Prevention Program (DPP) [4] and Finnish Diabetes Prevention Study (DPS) [5] on lifestyle modifications (low-fat diets, 5-7% weight loss, and exercise habits) have demonstrated a reduction in the burden of T2D by up to 58% [4]. A meta-analysis [17] of data from 11 randomized controlled studies (1369 participants) revealed that a low-carbohydrate diet can aid in weight reduction [18]. Moreover, a low-carbohydrate diet was also found to be more effective in glycemic control compared to a low-fat diet in patients with T2D [19].

While generalized dietary strategies such as low-fat and low-carbohydrate diets have been successful to varying degrees in various contexts, it is unclear whether and which approach may be successful for a specific individual. Advances in precision nutrition are promising but still under development and may not be cost-effective [10]. To address the need for individualized dietary recommendations, we explore the use of a computational simulation modeling tool in this work.

We previously developed a computational simulation model [20] of macronutrient metabolism and T2D onset and progression and tested it using data from DPP. The impact of lifestyle changes on endpoints including body weight and HbA1c were predicted at the individual level over a period of 3 years for 315 subjects from the DPP study. The mean prediction error for individual-level body weight and HbA1c changes over the three-year period was approximately 5% each. This suggests that the model can be used to predict and optimize individual-level responses to lifestyle changes. To our knowledge, currently there are no studies on the optimization of dietary strategies for preventing T2D using simulation modeling based on physiological principles.

The Japan Diabetes Outcome Intervention Trial-1 (J-DOIT1), a nationwide pragmatic cluster-randomized controlled trial, showed that participants who received telephone calls more frequently had a significantly reduced risk (41%) of T2D development [21,22]. Herein, using the simulation model, we analyzed data from J-DOIT1 [22] to evaluate factors affecting inter-individual variability in response to diet change, including endogenous (physiological characteristics) and exogenous (e.g., macronutrient intake) factors. The model adequately described individual-level body weight and HbA1c dynamics over time. We also demonstrate how the simulation approach may be used to optimize diet therapy for individuals to achieve specific health goals.

## Methods

### Simulation model

A previously developed computational simulation model of T2D was used [20]. This computational simulation model of T2D, referred to as the “model” henceforth, is based on the physiological mechanisms underlying the onset and progression of T2D. Important physiological (endogenous) and lifestyle (exogenous) factors involved in T2D are represented in the model. Exogenous factors influencing T2D are represented through dietary intake of macronutrients, i.e., carbohydrates, fats, and proteins, as well as energy expenditure through physical activity. Endogenous or physiological drivers of T2D are represented mechanistically in the model through physiological processes occurring at the cellular, tissue, and whole-body levels.

At a high level, the model mathematically represents the dynamics of dietary intake of carbohydrates, fats, and proteins, their breakdown and transportation into major tissue compartments through the bloodstream, and the interconversion of metabolic species into stored and active forms (**Error! Reference source not found.**). A module representing the pancreas regulates insulin secretion into the bloodstream. Cellular processes modulating the activation of insulin receptors by insulin drive the development of insulin resistance, which in turn controls several processes, including glucose uptake by tissues. Oxidation of macronutrients generates ATP, which provides energy for basal metabolism and physical activity. Changes in caloric intake, macronutrient composition, and/or physical activity levels have a cascading impact on all components of the model, leading to changes in key outputs, such as body weight, plasma glucose, and HbA1c.

Details regarding the development and validation of the model have been described previously [20]. For the analysis presented here, the model described in the original publication was used.

### Digital twins

The computational simulation model comprises several numerical parameters that can be adjusted to fit model outputs, such as body weight and HbA1c trends over time, to the observed data of a specific individual. A model that has been calibrated to represent the historical data of a specific individual can be considered a “digital twin” of the individual. The digital twin can be used to simulate experiments with various lifestyle modifications quickly and safely in a virtual *in silico* environment. The model’s ability to use digital twins to predict body weight and HbA1c was previously tested using individual-level data from DPP [4,20]. The concept of digital twins was applied in the work presented here. Digital twins were created for individuals selected from the J-DOIT1 study by calibrating instances of the model using a previously described method [20]. The digital twins were then used to simulate various scenarios to understand and analyze the variability in individual responses to interventions.

### J-DOIT1 study

The Japan Diabetes Outcome Intervention Trial-1 (J-DOIT1) is a pragmatic, cluster-randomized controlled trial conducted in Japan. The trial investigated the impact of lifestyle coaching delivered through telephone calls on T2D development in high-risk individuals in a primary healthcare setting [21]. A total of 2607 individuals (1240 in the intervention arm and 1367 in the control (placebo) arm) completed the study with a median follow-up period of 4.2 years [22]. Participants in the intervention arm received lifestyle support telephone calls from healthcare providers over a 1-year period. The intervention arm was further divided into three lifestyle support centers designated as centers A, B, and C. During the 1-year period for which telephone-delivered lifestyle support was provided, participants in centers A, B, and C received 3, 6, and 10 support calls, respectively. Thus, centers A, B, and C can be considered as low-, medium-, and high-support call frequency groups, respectively. The control arm did not receive any support through telephone but received periodic newsletters on diabetes and diabetes prevention. The participants were followed-up annually. The onset of T2D status was assessed as the primary outcome, and the other outcomes included body weight and HbA1c.

### Patient recruitment

Using the 2006 health checkup data, candidates who met the inclusion criteria were identified in each group. Inclusion criteria included an age of 20-65 years and impaired fasting glucose (IFG), defined as a fasting plasma glucose concentration (FPG) of 100-125 mg/dL (5.6-6.9 mmol/L). Exclusion criteria included diagnosed diabetes, a history of taking anti-diabetic agents, and a HbA1c of ≥ 6.5% [13]. Women with a history of gestational diabetes could be enrolled. Physical or medical conditions that do not allow exercise, pregnancy or possible pregnancy, type 1 diabetes mellitus, liver cirrhosis or chronic viral hepatitis, and the use of a cardiac pacemaker were also exclusion criteria. The study participants were registered from March 31, 2007, to January 25, 2008. The follow-up of the participants ended in March 2011.

### Selection of the analysis dataset

A total of 112 unique J-DOIT study participants were selected for the individual-level analysis using the following algorithm (**Error! Reference source not found.**). For each subject in the J-DOIT1 dataset, the percentage change in the body weight and HbA1c level from baseline to the end of the intervention was calculated. The degree of response for each subject was defined as the sum of the percentage decrease in the body weight and HbA1c. Individuals with the largest collective decrease in the body weight and HbA1c were considered as the “best responders” while those with the least decrease or greatest increase were considered as the “worst responders.” Using this definition, 29 best responders were selected from the intervention arm, with 10 each drawn from the low- and high-support call frequency groups, and 9 from the medium-support call frequency group (corresponding to centers A, C, and B, respectively, as described above). Similarly, 30 worst responders were selected from the intervention arm, with 10 each from the low-, medium-, and high-support call frequency groups. Thus, 59 subjects were selected from the intervention arm with nearly equal representation of the best and worst responders from all three call frequency groups.

Subsequently, a baseline-matched subject from the control arm was identified for each of the 59 subjects selected from the intervention arm. The method used to identify baseline-matched subjects is described next. Sex, height, baseline age, baseline body weight, and baseline HbA1c levels of each subject from the intervention arm were selected as the reference values. Matched subjects in the control arm with the same sex, height within ±3 cm, baseline age within ±2 years, baseline body weight within ±4 kg, and baseline HbA1c within ±0.3% of the reference value were selected. Of the subjects from the control arm that matched these criteria, the subject with the smallest difference in body weight and HbA1c level was selected as the baseline-matched pair of the intervention subject. If a matched subject from the control arm could not be found for a subject from the intervention arm, the intervention arm subject was dropped and another intervention subject was selected.

Using these criteria, 53 unique subjects were selected from the control arm. The number of unique subjects selected from the control arm was less than 59 because 6 control subjects were baseline-matched to 2 intervention subjects each. The 53 matched subjects from the control arm were used as the training dataset, and the other 59 from the intervention arm were used to test the model predictions. Further details of the training and test processes are described below. 12 unique subjects were selected for individual-level analysis. 59 subjects were selected from the intervention arm of J-DOIT1 with a nearly equal distribution over three call frequency groups and two response categories within each call frequency group. 53 subjects from the control arm were found to be the best baseline-matched pairs of the 59 subjects from the intervention arm.

### Model calibration and testing

The model consists of two types of parameters: 1) physiological parameters or parameters representing endogenous processes that are inherent to the individual and do not change over the course of the simulation; and 2) lifestyle parameters, which can change dynamically over time because of interventions.

### Calibration of the training dataset

For the training dataset, a subset of physiological parameters was calibrated in addition to lifestyle parameters (Table 1) to fit the model’s predicted body weight and HbA1c levels to each subject’s measured body weight and HbA1c time course over the duration of the J-DOIT1 study. While the physiological parameters were constant for an individual by design, step changes in lifestyle were allowed at discrete time points over the duration of the simulation. The set of physiological and lifestyle parameters that resulted in the best achievable fit to the measured body weight and HbA1c time course of an individual was accepted as the parameter set for that individual. As a result of this process, each subject from the training set had a unique combination of physiological and lifestyle parameters that defined the digital twin of that subject.

**Table 1.** Model parameters calibrated to fit individual subject time-course data.

| Category | Parameter fit to individual subject | Parameter symbol<sup>3</sup> |
| --- | --- | --- |
| Physiology parameters<sup>1</sup> | Basal carbohydrate intake requirement to maintain steady state body weight | $CI_0$ |
| | Basal fat intake requirement to maintain steady state body weight | $FI_0$ |
| | Maximal HbA1c concentration | $Cmax_{hba1c}^{BLD}$ |
| | Initial HbA1c concentration | $C_{hba1c}^{BLD}(t = 0)$ |
| | Maximal inhibitory effect of free fatty acids (FFA) on insulin signaling | $\alpha_{dep\_ffa}$ |
| | FFA concentration for half maximal inhibition of insulin signaling | $k_{dep\_ffa}$ |
| | Extent of pancreatic beta cell damage due to glucotoxicity, lipotoxicity, and inflammation | $\alpha_{bc,s\_ros}$ |
| Historical lifestyle parameters<sup>1</sup> | Carbohydrate intake prior to study start | $CI$ |
| | Fat intake prior to study start | $FI$ |
| Lifestyle parameters during the study<sup>2</sup> | Carbohydrate intake at various time points during the study | $CI_1, CI_2, ..., CI_4$ |
| | Fat intake at various time points during the study | $FI_1, FI_2, ..., FI_4$ |
| | Change in physical activity at various time points during the study | $\Delta PA_1, ..., \Delta PA_4$ |
| | Time points at which carbohydrate intake, fat intake, and physical activity change during the study | $T_1, T_2, ..., T_4$ |

<sup>1</sup>Calibrated only for baseline-matched subjects from the control group, i.e., training dataset.
<sup>2</sup>Calibrated for all subjects.
<sup>3</sup>Symbols as used in the original model [20].

### Calibration of the test dataset

As described above, the training dataset was obtained by baseline-matching the test data. The baseline-matched pairs comprising one subject each from the training and test datasets were of the same age and sex and had similar body weight, height, body mass index, and HbA1c at baseline. Because of this similarity in their baseline attributes, we assumed that the physiological parameters, as well as carbohydrate and fat intake prior to the start of the study were identical for both subjects in a baseline-matched pair. The implication of this assumption is that the physiological parameters of each test subject are predetermined by their corresponding match from the training dataset; any differences in the observed body weight and HbA1c time courses of the pair during the J-DOIT1 study could be explained only by differences in their lifestyles, such as carbohydrate and fat intake and exercise changes during the study. This limits the range of responses that can be achieved for individuals in the test dataset because lifestyle is the only variable input to the model and serves as a mechanism to test the model’s ability to forecast individual responses. For the test dataset, only step changes in the category “Lifestyle parameters during the study” (Table 1) were allowed. The time points at which these step changes in lifestyle were introduced in the simulation were determined empirically based on trends in body weight and HbA1c. Whenever a previously decreasing trend in either body weight or HbA1c was followed by an increasing trend or vice-versa, a lifestyle change was introduced, assuming that such changes in body weight or HbA1c could only be driven by lifestyle factors. A maximum of four such discrete lifestyle changes were permitted for each subject over the approximately 4-year follow-up. Changes in lifestyle parameters were calibrated for each test subject to determine the best fit to individual time courses of body weight and duration over the duration of the J-DOIT1 study.

Parameter calibrations were performed using the differential evolution algorithm [23] and the objective function to be minimized was the sum of the squared errors over all time points for body weight and HbA1c. For calibration, each data point was assumed to have an inherent measurement error, and the objective function was designed to consider this error. Body weight was assumed to carry a measurement error of ±1 kg based on previous studies on imprecision in the measurement of body weight using weighing scales [24,25]. HbA1c was assumed to have a measurement error of ±0.15 percentage points, which is approximately 3% of the median HbA1c value of 5.5% across all data points in this analysis. A 3% error is well within the ±5% measurement error considered acceptable by the National Glycohemoglobin Standardization Program (NGSP) [26]. Based on a study of Japanese individuals, the measurement error for HbA1c was estimated to be 0.17 percentage points [27].

The following objective function was used for parameter estimation for each subject:

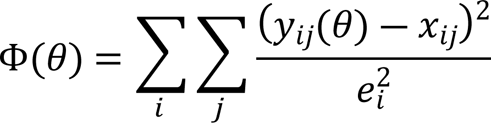

where 𝜃 represents the model parameter vector, 𝑖 is either body weight or HbA1c and 𝑗 represents all the time points at which biomarker 𝑖 is measured for the subject. 𝑒_𝑖_is the measurement error associated with biomarker 𝑖, such that 𝑒_𝑏𝑜𝑑𝑦 𝑤𝑒𝑖𝑔ℎ𝑡_ = 1 𝑘𝑔 and 𝑒_𝐻𝑏𝐴1𝑐_ = 0.15%.

### Simulations

To test the effects of dietary changes and determine the optimal diet, simulations were performed using the calibrated digital twins of the study subjects. Starting from the baseline age (age at the start of J-DOIT1) of a digital twin, a random step change in carbohydrate and fat intake was introduced. Keeping all other parameters constant, the body weight and HbA1c time-courses were simulated with diet change. This process was repeated multiple times for each digital twin using a Monte Carlo approach with macronutrient changes sampled from a uniform random distribution in the range baseline value – 25% to baseline value + 25%. The simulation outputs were recorded and analyzed.

## Results

### The model successfully captures individual-level dynamics of body weight and HbA1c

The model was fit to individual time-courses of body weight and HbA1c by calibrating both physiological and lifestyle parameters (Table 1) for the training dataset and only lifestyle parameters for the test dataset, as described in the Methods section. Results showed that individual-level changes in the body weight and HbA1c over time were captured well by the model for both the training and test datasets (Fig 3,**Error! Reference source not found.** Supplementary Fig S1-S6). Visual comparison of the predicted values with the measured values across all time points for all subjects indicated that the model performs well at predicting the measured values (Supplementary Fig S7). The prediction error (mean [±SD]) across all data points in the training dataset for body weight was 0.7 kg (±0.8) and for HbA1c it was 0.08% (±0.08). In terms of percentage error (mean [±SD]), body weight of subjects in the test dataset was predicted with an error of 1.1% (±1.0) and HbA1c with an error of 1.4% (±1.4) relative to the actual measurement (Table 2).

**Figure 1.**
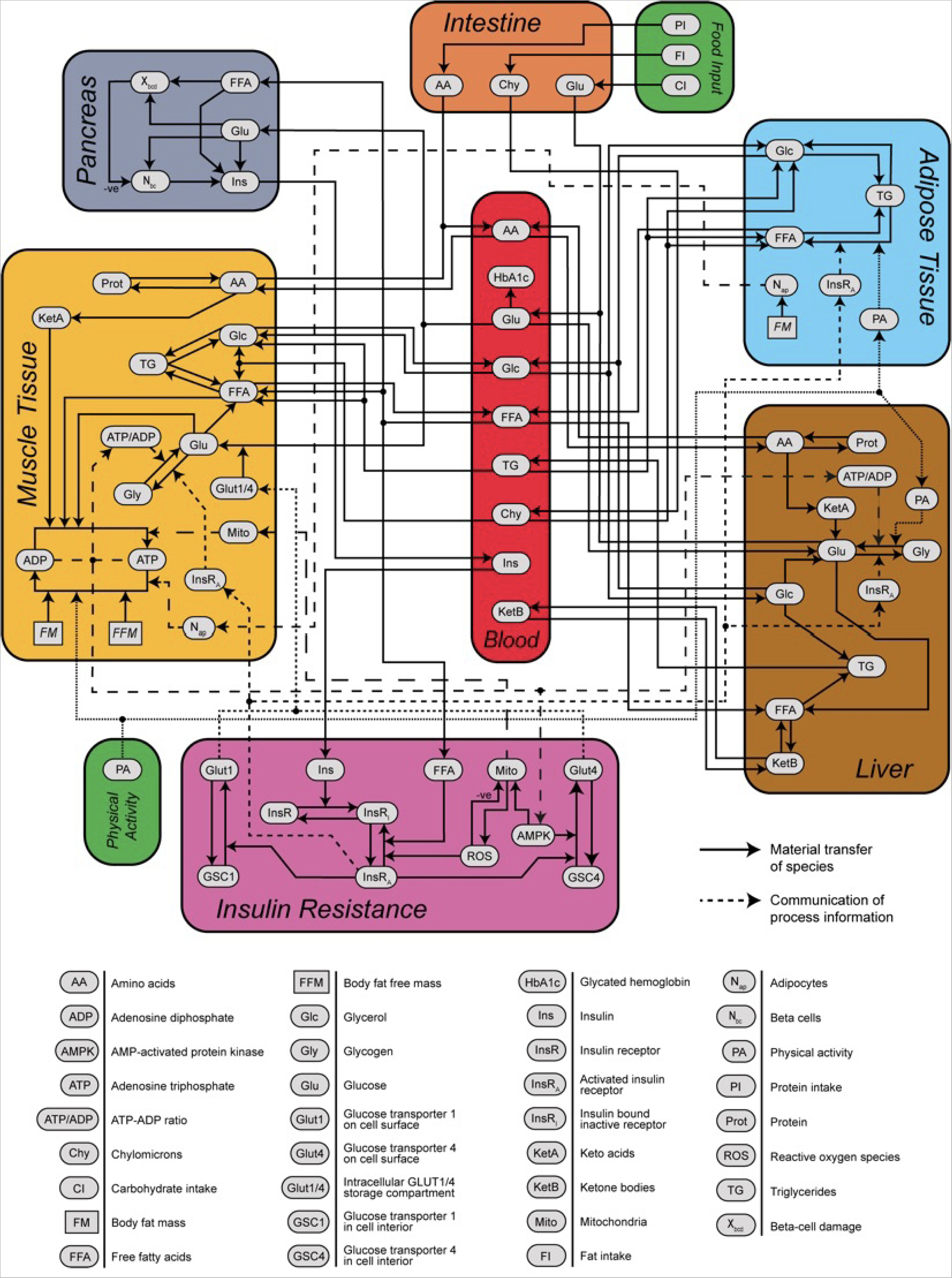
Simulation Model.

**Figure 2.**
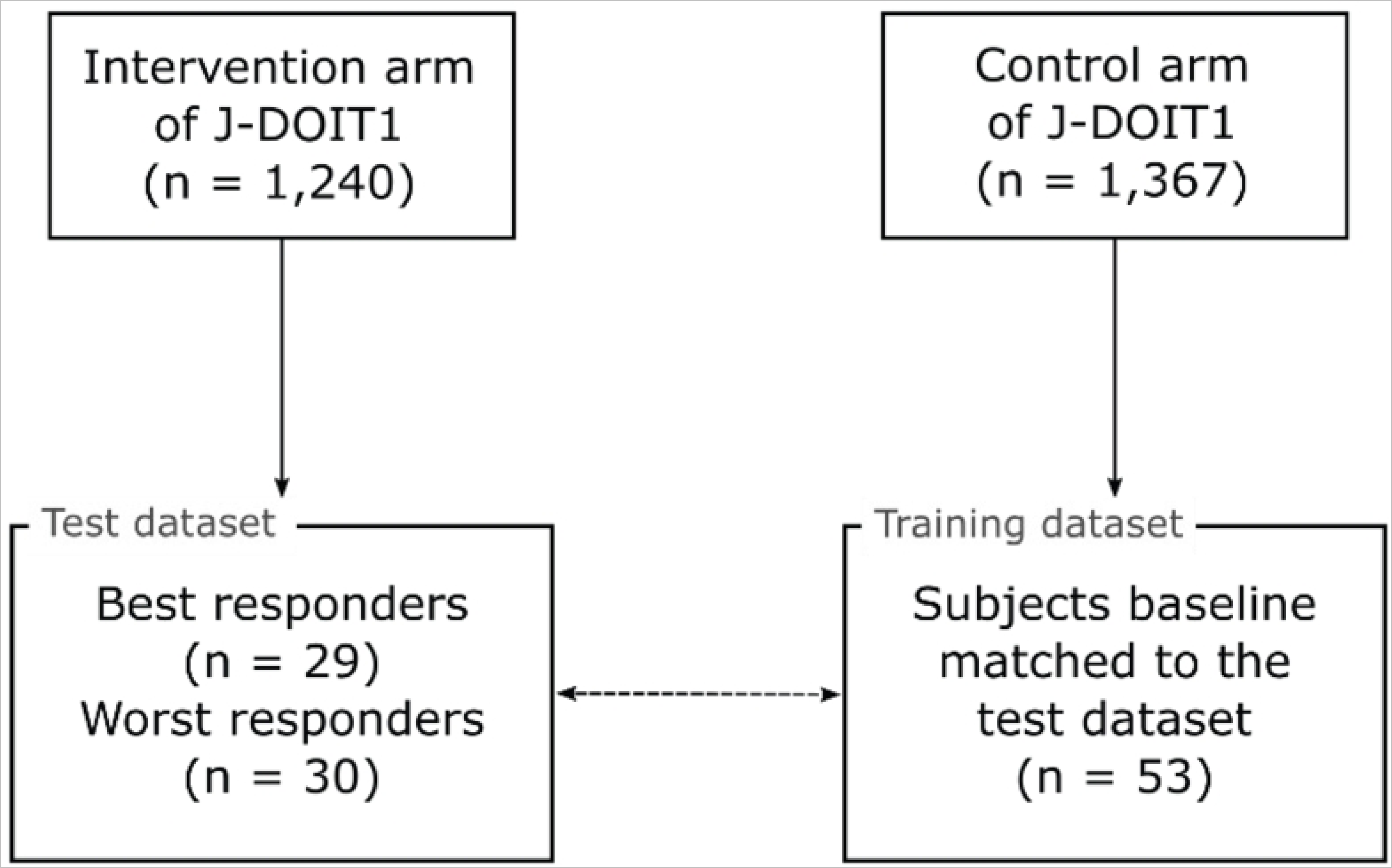
Subject selection and study design.

**Figure 3.**
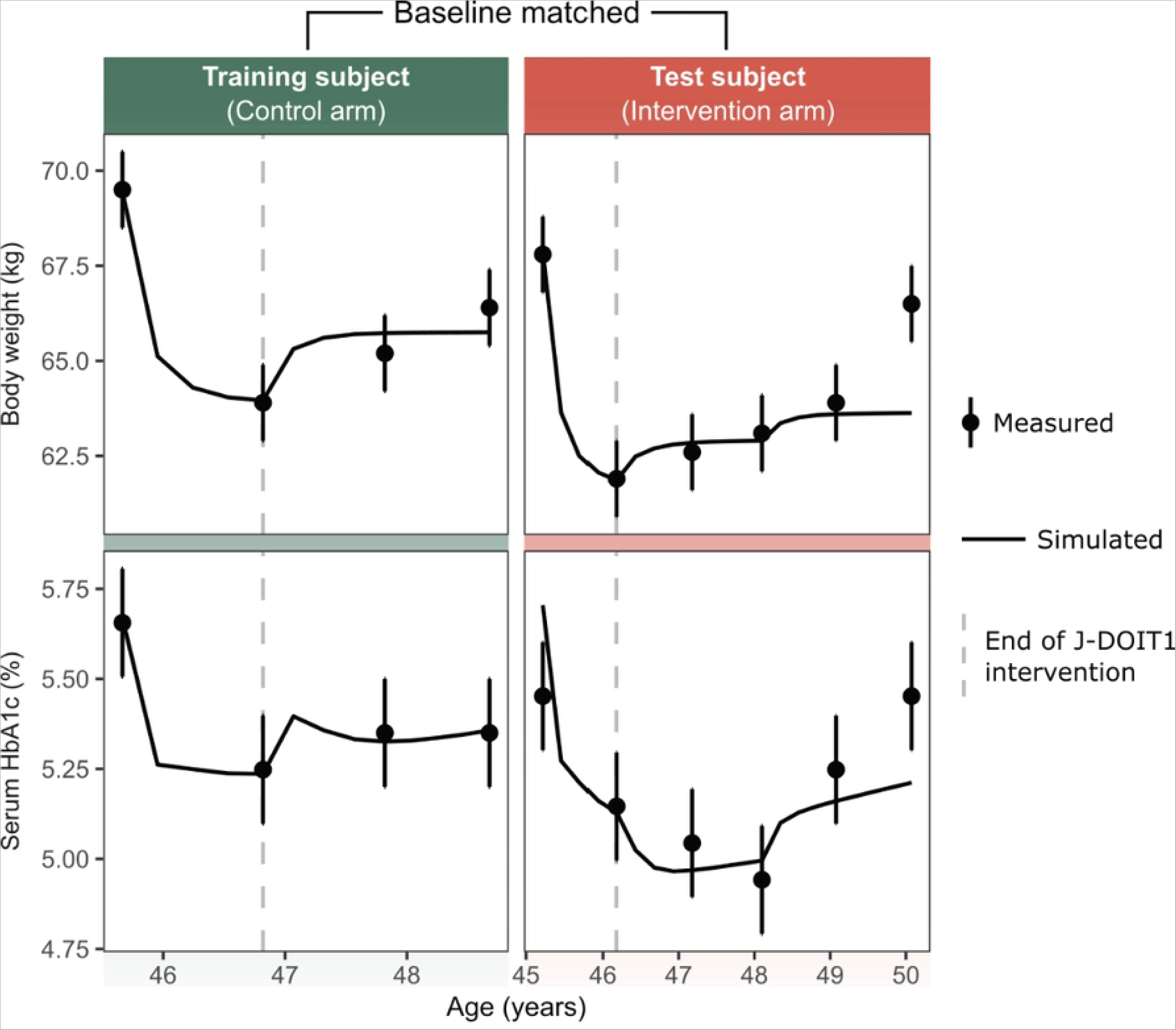
An example of model prediction for a pair of baseline-r

**Table 2.**
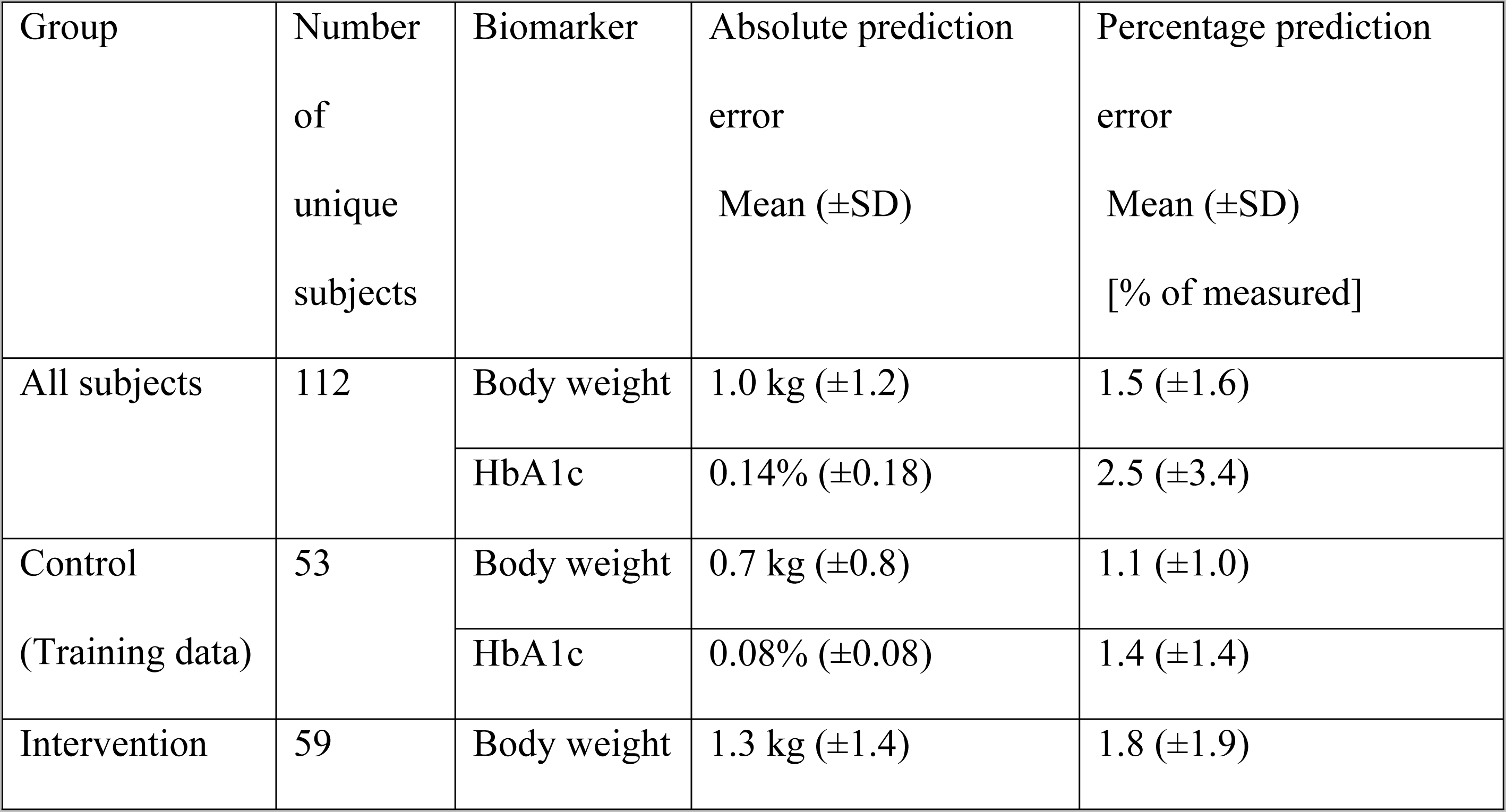
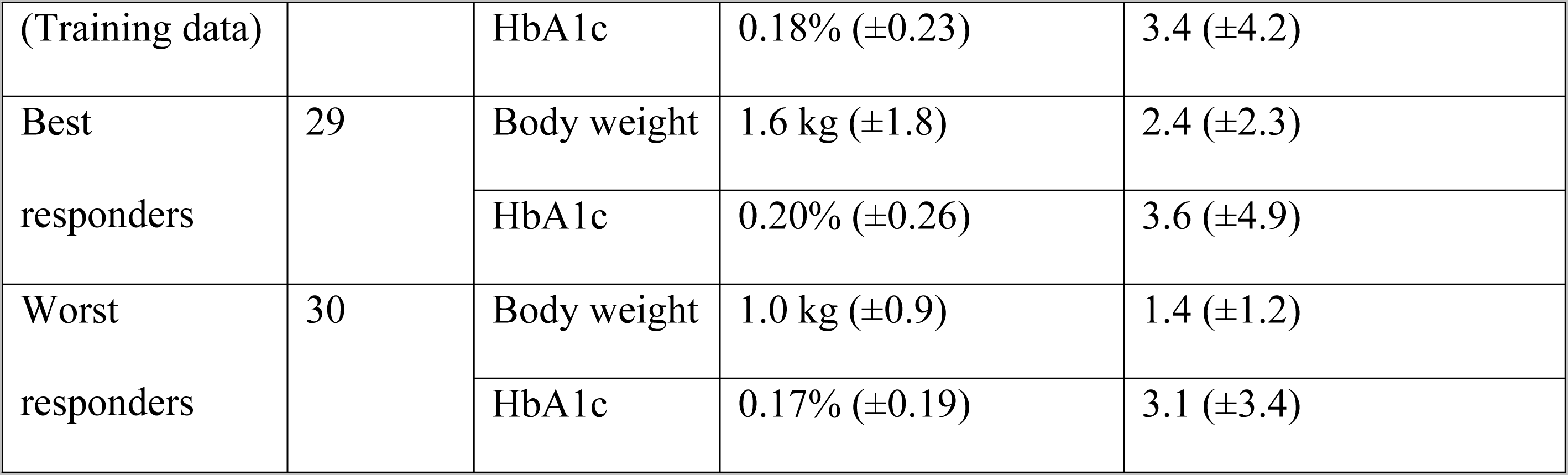
Model prediction errors. Prediction errors are shown after grouping subjects using 306 various criteria.

Panels on the left-hand side represent a subject from the training data set. Panels on the right-hand side show the baseline-matched subject from the test data set. The test subject is from the high call frequency group and was classified in the best responder category. The error bars around the measured values are assumed measurement errors, ±1 kg for body weight and ±0.15 points for HbA1c, as described under model calibration in the Methods section

The prediction error (mean [±SD]) across all data points in the test dataset for body weight was 1.3 kg (±1.4), and for HbA1c it was 0.18% (±0.23). In terms of percentage error, body weight was predicted for the test dataset with an error of 1.8% (±1.9) and HbA1c with an error of 3.4% (±4.2) relative to the measured value (Table 2).

### Changes in caloric balance alone do not fully explain the variability in individual response

After calibration and testing against individual time-course data, the model was used to estimate the likely caloric change per individual that led to the observed change in body weight. Calibrated digital twins were used to estimate the caloric change for each individual due to modifications in diet and exercise during the period between baseline and first follow-up in the intervention period of the J-DOIT1 study (median duration 1 year). The total caloric change (decrease or increase) was defined as the sum of changes in caloric intake due to diet change and caloric expenditure due to exercise. Changes in daily calories from baseline to the first post-baseline follow-up were estimated for each individual using the calibrated model parameters. The measured change in body weight during the same interval (baseline to the first follow-up) was also calculated. The model-estimated caloric change versus the observed weight change from baseline to the first follow-up is shown in Fig 4 **Error! Reference source not found.**. The measured change in body weight generally increased with the model-predicted increase in caloric intake, with a Pearson correlation coefficient of 0.82 (**Error! Reference source not found.**). The model predicted that similar caloric changes could lead to a wide range of responses in terms of body weight changes across individuals, as indicated by the spread of the points along the y-axis in **Error! Reference source not found.**. When a linear regression model was fit to the data (solid gray line in **Error! Reference source not found.**), the residual error ranged from −4.6 kg to +7.0 kg with a residual standard error of 2.5 kg, indicating a relatively wide spread of body weights around the line of best fit. This suggests that changes in calorie intake alone may not be sufficient to predict individual-level changes in body weight. Similar trends were observed for HbA1c levels (Supplementary Fig S8).

**Figure 4.**
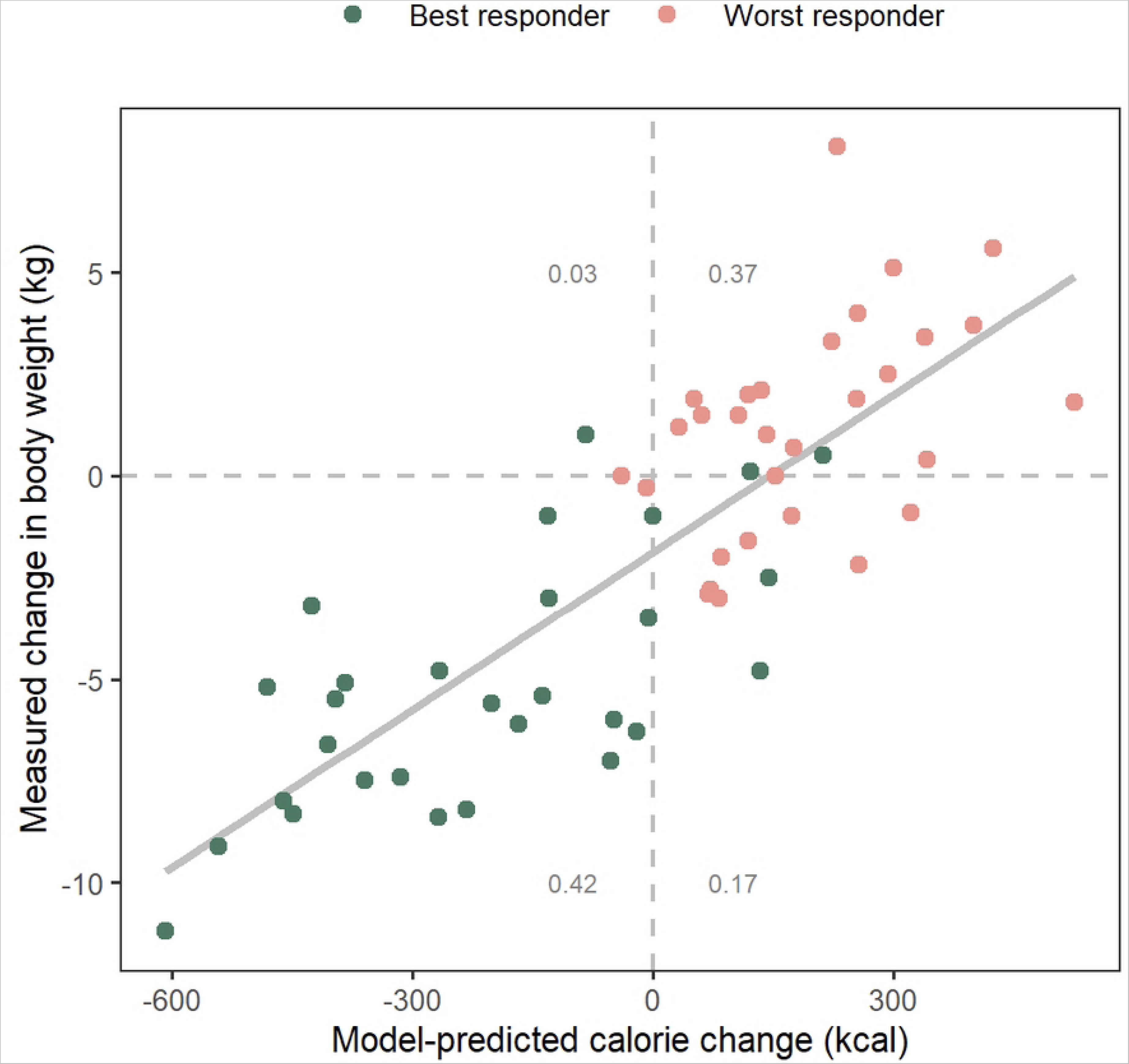
Model-predicted caloric change versus weight change

The measured change in body weight from baseline to the first follow-up during the J-DOIT1 intervention (median duration 1 year) is plotted against model-estimated change in calories per day due to both diet and exercise changes averaged over the same period for subjects in the intervention arm. The gray number in each quadrant is the fraction of data points in that quadrant. The data points fit a linear regression model (solid gray line) with r^2^ = 0.67 and a residual standard error of 2.5 kg, indicating a relatively widespread around the line of best fit. Best and worst responders were defined based on the total percent change in body weight and HbA1c one year after the end of the J-DOIT1 intervention.

We also explored the question of whether the degree of response (change in body weight and HbA1c) could be related to endogenous characteristics (physiology parameters defined in Table 1) of subjects. None of the calibrated physiology parameters, either alone or in linear combinations, were found to be correlated with changes in body weight or HbA1c.

### Diet therapy is predicted to have maximal effectiveness when optimized individually

Simulations were performed to determine the “optimal” diet for achieving a 5-7% reduction in body weight over a period corresponding to the duration between baseline and 1-year post-intervention. Digital twins of the J-DOIT1 study subjects from the test dataset (N = 59) were simulated with various random modifications to their carbohydrate, fat, and protein intake. Each macronutrient was sampled from a uniform distribution within ±25% of its <u>baseline</u> value for the digital twin. Diets that led to a 5-7% reduction in body weight were selected as optimal diets. Using this approach, optimal diets could be identified for 48 of the 59 subjects; the remaining 11 subjects probably needed diet changes beyond the ±25% range simulated. Of the 48 subjects for whom optimal diets could be identified, only a single diet change (24% reduction in carbohydrate and 25% reduction in fat intake) led to a 5-7% reduction in body weight.For all other subjects (N = 47), there was no single optimal diet that led to a 5-7% reduction in body weight; instead, a set of various diet compositions could lead to the target weight reduction (a range of 3 to 668 diet compositions for each subject with a median of 186 diet compositions). Furthermore, this set of diets was unique to each participant. A comparison of distributions of the optimal diets for two subjects is shown as an example in Fig 5 **Error! Reference source not found.**.

**Figure 5.**
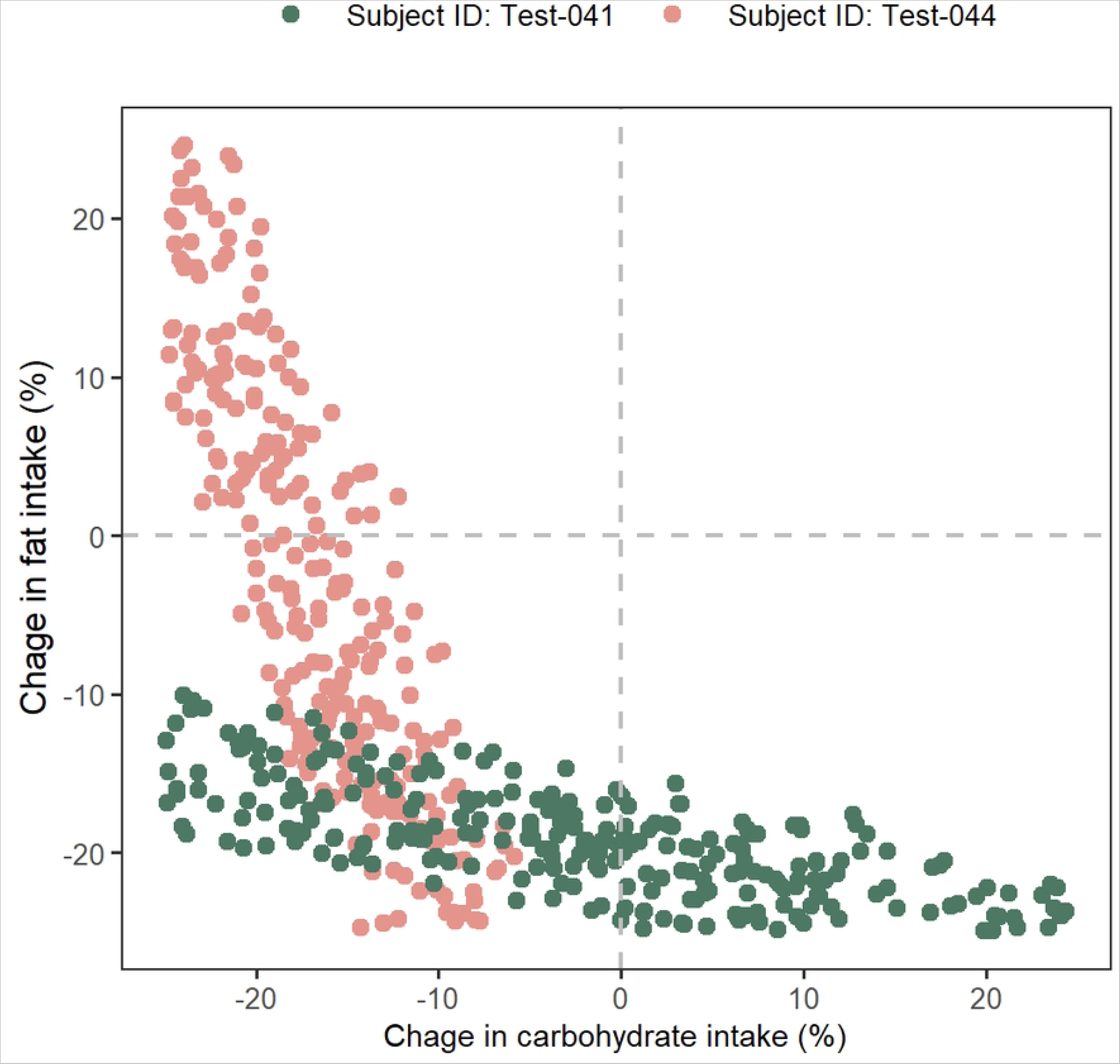
Optimal changes in carbohydrate and fat intake for targ

The two subjects presented in Fig 5 **Error! Reference source not found.** show qualitatively different distributions of optimal diet changes. For subject ID Test-041, carbohydrate intake could change over a wide range of approximately −25% to +25% but fat change needed to be more narrowly restricted between approximately −25% to −10%. Contrary to this, for subject ID Test-044, fat change could range between −25% to +25% but carbohydrate change had to be restricted to a narrower range (−25% to −5%). In an alternative interpretation, subject ID Test-041 is predicted to be more sensitive to fat change than to carbohydrate change and should more precisely control fat intake to achieve the targeted weight loss. Subject ID Test-044, on the other hand, is predicted to be more sensitive to carbohydrate change; this subject should pay more attention to regulating carbohydrate intake but can be less particular about controlling fat intake. Monte Carlo simulations identified a unique set of “optimal” carbohydrate and fat changes required for each subject that were predicted to lead to a targeted 5-7% reduction in body weight.

In addition to the 5-7% body weight reduction for subject Test-041(Fig 5), an additional target of 0.1-0.2 point reduction in HbA1c was added. Applying this additional target led to further refinement of the optimal diets and a subset of the original optimal diets was predicted to simultaneously achieve both targets (Fig 6).

**Figure 6.**
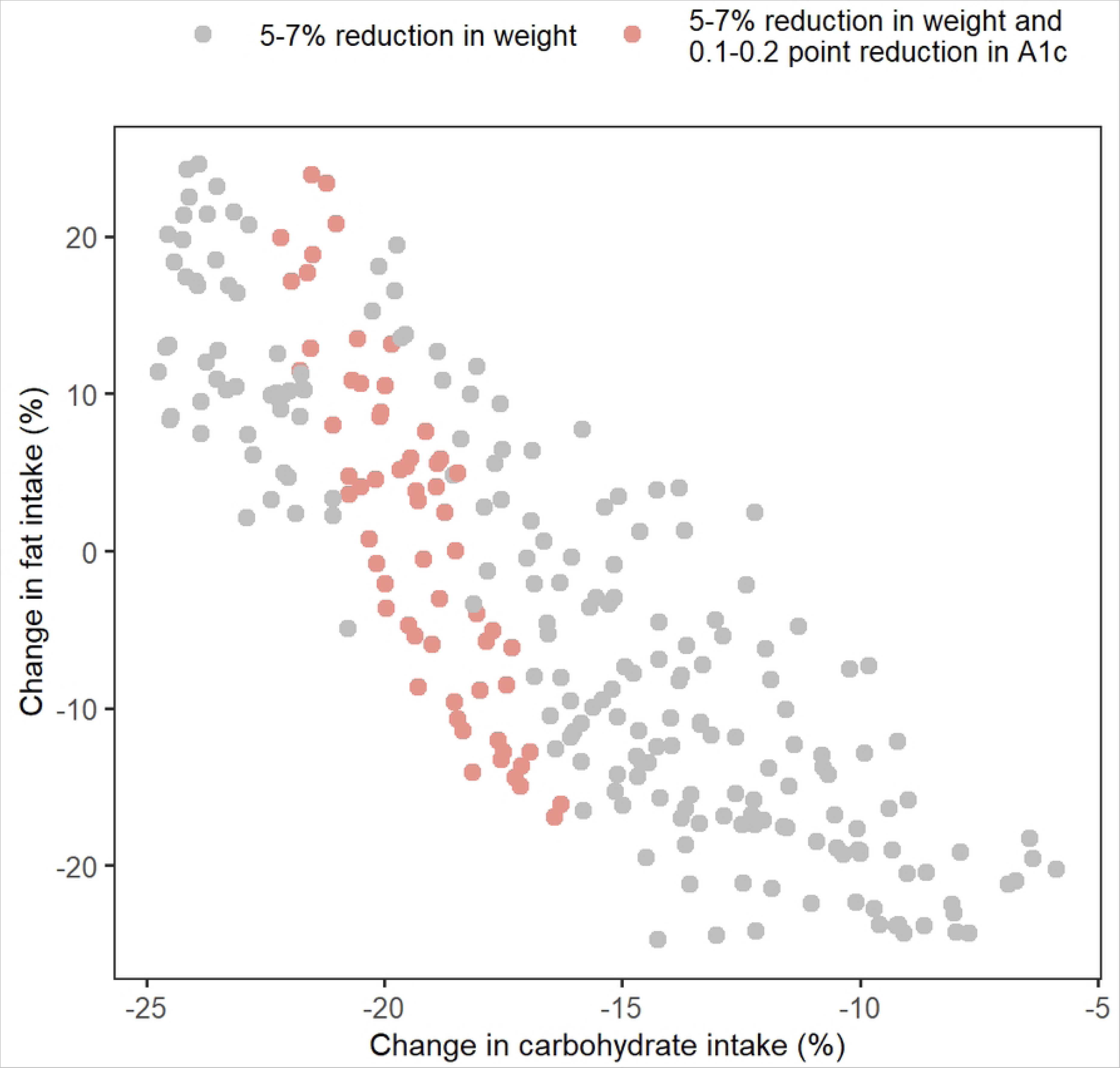
Including additional biomaker targets furthers narrows

The subset (pink circles) of optimal diets identified for subject Test-041 (Fig 5) to achieve a 5-7% reduction in body weight (gray and pink circles) was predicted to additionally reduce HbA1c by 0.1-0.2%.

### Individuals show differential sensitivity to carbohydrate and fat changes

The simulation-based diet optimization results were used to explore whether all subjects could be classified into carbohydrate or fat sensitive categories. After finding the set of optimal diets for each subject using simulations as described above, lines of best fit were obtained for each subject’s (N = 47 subjects with >1 optimal diets) predicted set of optimal diet changes (Fig 7). These lines approximate the predicted optimal diet change patterns for each subject and are a reasonable simplification for easy visualization and analysis of the diet patterns. All lines had negative slopes implying that if a subject were to shift to a smaller reduction in carbohydrate intake, it could be compensated by a larger reduction in fat intake, and vice versa. Additionally, the shifts would have to move along the line, so the magnitude of compensation required was different for each subject as determined by the slope of the line.

**Figure 7.**
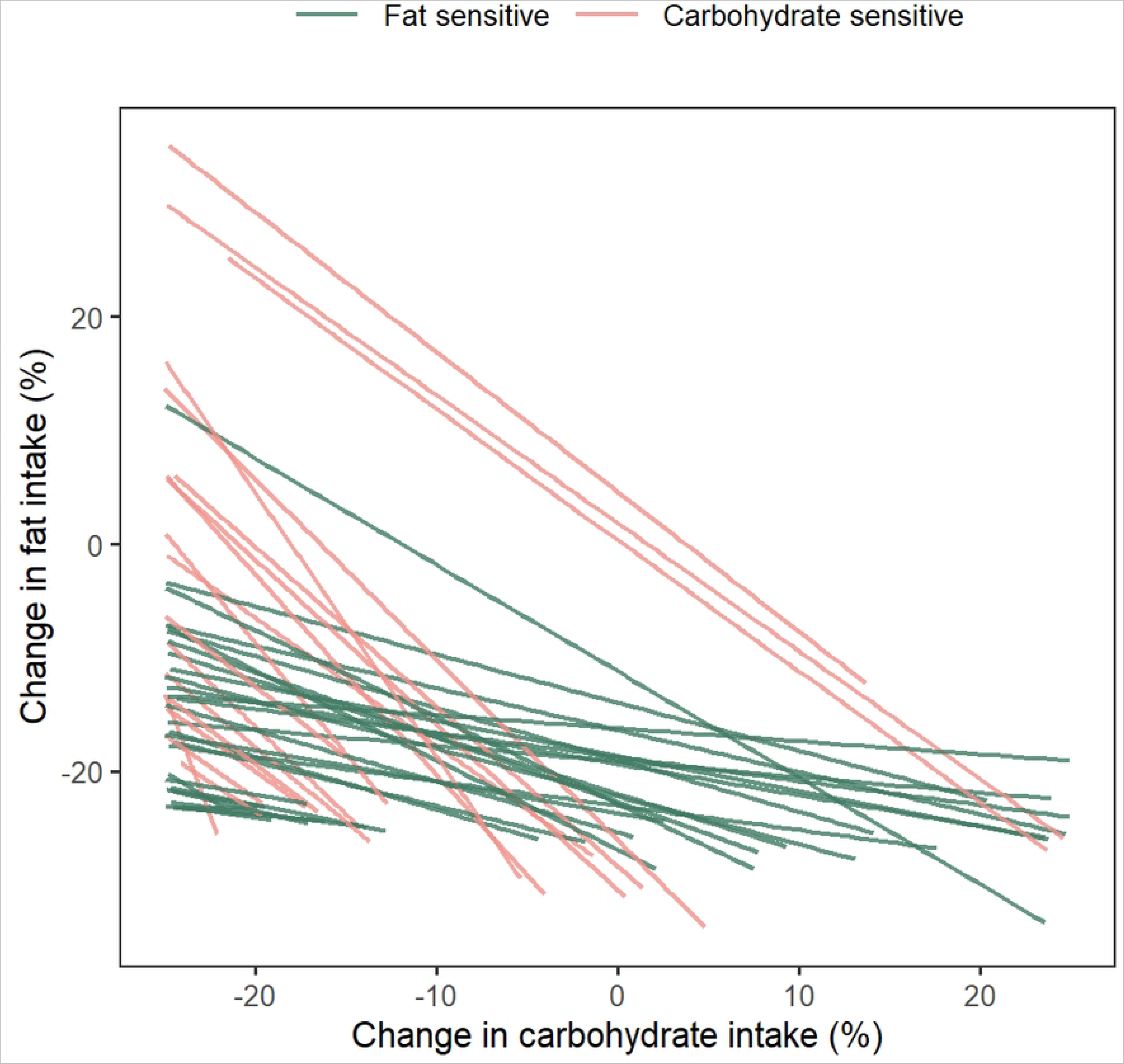
Optimal diet trajectories and relative sensitivity to mac

For a hypothetical subject whose line of best fit has slope of −1 (angle of −45° with the x-axis), a downward (upward) shift of X% in carbohydrate change could be compensated by a corresponding upward (downward) shift of exactly X% in fat change. Therefore, a subject with a slope of exactly −1 can be considered to be equally sensitive to changes in carbohydrate and fat intake. As the line becomes increasingly horizontal (angle with the x-axis between −45° and 0°, slope between −1 and 0), the sensitivity regime shifts towards greater sensitivity to fat change because for a nearly horizontal line, fat change must be tightly controlled while carbohydrate change can vary widely. Conversely, as the line becomes more vertical (angle with the x-axis between −90° and −45°, slope < −1), it indicates a greater sensitivity to carbohydrate change. Based on these concepts, individuals were classified as carbohydrate sensitive (slope < −1) or fat sensitive based on the slopes of their lines (slope > −1) (Fig 7). A total of 29 (62%) subjects were identified as has having a greater sensitivity to fat change and 18 (38%) as being more sensitive to carbohydrate changes based on the sensitivity criteria defined above.

A line was fit to the set of optimal diets predicted for each subject. The slopes of the lines were used to classify subjects into carbohydrate or fat sensitive categories. Lines that tend to be more horizontal (green lines; slope > −1) indicate individuals with greater sensitivity to fat change. Lines that tend to be more vertical (pink lines; slope < −1) indicate individuals with greater sensitivity to carbohydrate change.

## Discussion

Diet therapy can be an effective non-pharmacological method to delay or prevent the onset of T2D; however, diet therapy has not been shown to be consistently effective [4–6,22]. The lack of effectiveness of diet therapy could be due to personalized dietary requirements [7–9]. Previous studies showed that individuals receiving an identical standardized low-energy diet show variability in their weight trajectories [28]. Metabolic heterogeneity among individuals could be due to genetic and epigenetic factors, microbiome, lifestyle, and environmental exposure [29]. Personalized nutrition is a growing area of focus for both patients and experts. Optimizing diet change to individual physiological responses could maximize the impact of lifestyle intervention; however, tools that can enable customization of interventions at the individual level are lacking. We demonstrate, for the first time to our knowledge, the application of a computational simulation model as a tool to optimize diets for prediabetic individuals.

The emergence of digital twins and digital representation of objects or individuals provides a new opportunity to tailor individualized interventions [30]. We used a previously developed and tested mechanistic simulation model of human physiological processes involved in the onset and progression of diabetes to create digital twins of a subset of pre-diabetic subjects from the J-DOIT1 study. In the default setting, the parameters of the model are calibrated to represent a “typical” individual. When individual-level time-course data, such as body weight and HbA1c level over time are available, selected parameters of the model can be calibrated to fit the model to an individual subject’s data, which leads to a model customized to the subject, i.e., a digital twin of the individual. The digital twin provides a platform to conduct computational experiments quickly and safely in an *in silico* environment. Digital twins were utilized in this study to explore and optimize lifestyle recommendations through simulation.

We leveraged the simulation model to understand the inter-individual variability in responses to lifestyle interventions in the J-DOIT1 study. The selected individuals from the intervention arm were baseline matched with the participants from the control arm of the study. The baseline-matched individuals from the control arm formed the training set (n = 53), and individuals selected from the intervention arm comprised the test dataset (n = 59). Each subject from the training set was calibrated using the simulation model to generate a unique combination of physiological and lifestyle parameters that defined the digital twin of that subject. We assumed that individuals with similar baseline characteristics (age, sex, height, weight, and HbA1c) have similar physiological parameters and historical lifestyle. Therefore, physiological, and historical lifestyle parameters were replicated for the test subjects within each baseline-matched train-test pair. The post-baseline lifestyle parameters of the test subjects were allowed to change to explain the observed body weight and HbA1c trends. The digital twins thus generated captured the individual-level dynamics of the body weight with an error of 1.1% (±1.0) and HbA1c levels with an error of 1.4% (±1.4) relative to the actual measurements over a follow-up period of approximately 4 years.

The digital twins enabled the exploration of inter-individual variability in response to diet intervention. The digital twins created using the model were used to estimate the actual lifestyle change in terms of total caloric intake for all individuals in the training and testing datasets. We observed that the measured change in body weight generally increased with the model-predicted increase in daily caloric intake; however, similar changes in daily calories were predicted to result in a relatively wide band of weight change (approximately ±2.5 kg around the line of best fit, or a range of 5 kg) (Fig 4). This suggests that changes in calories alone may not be sufficient to predict individual-level changes in body weight and elucidates the significance of physiology, among other factors, in determining an individual’s response to diet.

Just as exogenous lifestyle factors were not fully predictive of the outcome, endogenous physiological parameters were also not found to be correlated with the outcome. This suggests that the outcome of a lifestyle change is an emergent property of complex interactions between underlying physiological processes and exogenous changes. Predicting such a response, therefore, requires an understanding of the complex interactions driving the response. Our physiology-based, quantitative framework, which captures such interactions by design, is well-suited for this purpose.

Having tested the model’s ability to satisfactorily describe individual-level dynamics of body weight and HbA1c, we applied the model to generate optimal diet recommendations for individuals in the training and testing datasets. Monte Carlo simulations were performed for each individual using their digital twin, and a unique set of “optimal” carbohydrate and fat changes required for a targeted 5-7% reduction in body weight was determined. The model predicted that there is no single optimal diet to achieve the target body weight. Analysis of optimal diet trajectories at the subject levels suggested that while some patients required tight control over fat intake (individuals sensitive to fat change), others required a a greater focus on managing carbohydrate intake (individual more sensitive to carbohydrate change). Furthermore, the set of optimal diets could be further refined by including additional goals, e.g., a targeted reduction in HbA1c. This result supports the role of personalized nutrition and dietary recommendations in improving health outcomes and demonstrates the potential utility of our approach in identifying such personalized recommendations based on historical subject data.

The modeling and analyses presented in this work are affected by a few limitations of data and methodology that should be acknowledged. The target population of our analysis only included Japanese individuals with prediabetes, thus limiting the generalizability of the predictions. The matching algorithm used to create pairs of train-test subjects allowed a small degree of mismatch so that matched pairs could be practically found. The assumption of physiological identity between the matched pairs has, therefore, some inaccuracy inherent to it and could impact the estimation of parameters as well as model predictions. Furthermore, all lifestyle changes were simulated as step functions, as this was mathematically the simplest form in the absence of additional information on individual lifestyle habits. In real life, lifestyle factors may be much more variable and may follow trends very different from a step function. This assumption is likely to impact the timing and rate of change of model-predicted variables like body weight. Finally, the mechanistic mathematical model used in this study makes several assumptions about the physiological processes underlying diabetes onset and progression, which may not always reflect the underlying biology and physiology accurately. Nonetheless, even with these limitations, the model predicted the body weight and HbA1c time courses of the training as well as test groups with high accuracy, which lends credence to the model and supports its use for predictive analysis.

An advantage of the model-based framework developed in this study over approaches like precision nutrition is that it can provide optimal dietary recommendations without requiring specific genetic and microbiome data, making it a quicker, lower-cost alternative. Prior validation of the simulation model using long-term data [20] and additional validation in this work using a subset of participants from the J-DOIT1 study showed that the framework predicts weight changes and glycemic control in individuals with high accuracy. This provides assurance that the framework can be used to predict optimal dietary recommendations for prediabetic individuals. Generatability was limited and careful attention should be paid for interpretation results because of the target population (Japanese adults with prediabetes). A prospective study in human subjects is required to build further confidence in this simulation model framework and confirm its utility in clinical practice.

The latest Dietary Guidelines for Americans (DGA) focus on limiting fat, especially saturated fat, and allowing higher carbohydrate intake. Volek *et al.* have argued that the DGA recommendations of a low-fat high-carbohydrate diet for the past several years have coincided with rapidly escalating epidemics of obesity and T2D that contribute to the progression of cardiovascular diseases [31]. This guideline lacks flexibility and does not appreciate the heterogeneity in individuals’ responses to dietary interventions. The findings of the J-DOIT1 study, coupled with the model-based framework for diet optimization presented in the study, offer additional evidence to convince experts and policymakers of the need for optimal diet interventions because of inter-individual variability in responses to identical diets. Our modeling framework can simulate changes in body weight and glycemic control as a result of lifestyle interventions at an individual level. The ability to optimize nutritional strategies using this model could help dieticians and physicians personalize diet recommendations to their patients’ goals.

## Disclosure of ethical statements

This study conformed to the standards of the Declaration of Helsinki。

Approval of the research protocol: The present study was approved by the ethics committee of the National Hospital Organization Kyoto Medical Center (No.16-102).

Informed consent or substitute for it was obtained from all patients for being included in the study.

Approval date of Registry and the Registration No. of the study/trial: Trial registration number: University hospital Medical Information Network (UMIN) Center: UMIN000000662)).

## Data Availability

Individual patient data in this study was obtained from the JDOIT-1 Study with permission from the study group. Queries related to the patient data should be directed to the JDOIT-1 study group. The simulation model used in this study was previously published - https://doi.org/10.1371/journal.pone.0192472

## Supporting information

**S1 Fig. Calibrated pairs for best responders, low call frequency group.**

**S2 Fig. Calibrated pairs for best responders, medium call frequency group.**

**S3 Fig. Calibrated pairs for best responders, high call frequency group.**

**S4 Fig. Calibrated pairs for worst responders, low call frequency group.**

**S5 Fig. Calibrated pairs for worst responders, medium call frequency group.**

**S6 Fig. Calibrated pairs for worst responders, high call frequency group.**

**S7 Fig. Measured vs Predicted Biomarkers.** Model-predicted body weight and HbA1c values for all subjects across time points show reasonable concordance with corresponding measured values with most values lying on or close to the line of identity.

**S8 Fig. Model-estimated change in calories vs measured change in HbA1c from baseline.** The measured change in HbA1c from baseline to the first follow-up during the J-DOIT1 intervention plotted against model-estimated change in calories per day due to both diet and exercise changes averaged over the same period for subjects in the intervention arm. The gray number in each quadrant is the fraction of data points in that quadrant. The data points fit a linear regression model (solid gray line) with r^2^ = 0.20 and a residual standard error of 0.28 points.

